# Implementation of the PrAISED (Promoting Activity, Independence and Stability in Early Dementia) intervention in practice: an evaluation using the Consolidated Framework for Implementation Research

**DOI:** 10.1101/2023.05.24.23289730

**Authors:** Emma J Adams, Clare Burgon, Juliette Lock, Helen Smith, Robert Vickers, Rachael Tucker, Stephen Timmons, Elizabeth Orton, Sarah E Goldberg, John Gladman, Tahir Masud, Rowan H Harwood

**Author notes:** Corresponding author: Emma J Adams.

## Abstract

**Background:** There is a paucity of evidence relating to the implementation of interventions for dementia care. The Promoting Activity, Independence and Stability in Early Dementia (PrAISED) intervention is a 12-month, home-based, individually tailored rehabilitation programme, delivered by therapists and rehabilitation support workers, with a focus on strength, balance, physical activity and activities of daily living which has been tested in a randomised controlled trial (RCT). The aim of this study was to identify what is required to implement PrAISED, or similar interventions, in a real-world setting in routine practice.

**Methods:** A 6-month version of PrAISED was delivered as a pilot service in one National Health Service organisation in England. Adaptations were made to intervention processes to facilitate the delivery of PrAISED as a service instead of as part of a research study. The number and duration of visits for each patient were recorded by intervention delivery staff and were summarised using descriptive statistics. Semi-structured interviews were conducted with seven members of staff delivering the PrAISED pilot service (two managers, five delivery staff) and eight members of staff from other sites involved in the PrAISED RCT (four managers, four delivery staff). The Consolidated Framework for Implementation Research was used to inform interview guides and conduct a codebook thematic analysis.

**Results:** Between April and November 2022, 11 patients were referred to, and participated in, the service. Patients received on average 20.9 visits (mean duration 82.1 mins). Five themes were identified from interviews relating to the pilot service: operational processes; workforce capacity; referral; intervention delivery and patient impact. A further six themes were identified regarding the wider implementation of dementia therapy programmes: the need for support post-dementia diagnosis; acceptability; effective delivery; reach/referral; intervention design and adaptability; and intervention materials and training.

**Conclusions:** Interventions like PrAISED are needed to fill a gap in support immediately post-dementia diagnosis. Future implementation in practice will require attention to the identification of intervention funding; leadership and management; time to establish operational processes; therapists with appropriate skills and experience; providing training and resources to support intervention delivery; defining patient eligibility, refining referral processes; and maintaining fidelity of the intervention.

## Background

Globally, more than 55 million people have dementia which will rise to over 150 million by 2050 [1, 2]. In the United Kingdom (UK), over 900,000 people are living with dementia [3] and the total cost of dementia care is estimated to be £34.7 billion [4], placing a significant burden on the economy and health and social care services. Dementia is a neurodegenerative condition associated with loss of memory and executive function, and changes in behaviour and mood [5]. Mild Cognitive Impairment (MCI) is also a syndrome of impaired cognition, with retained everyday functioning, which presents a significant risk for progression to dementia [6]. Impaired functioning in dementia may be a result of cognitive impairment, but also physical impairment such as abnormal or impaired gait [7, 8], or co-morbidities. Exercise and activity interventions involving strength and balance training and functional tasks, in particular dual-tasking (which involves simultaneously performing two or more tasks, such as a motor task and a cognitive task), may improve physical ability, balance and everyday functioning [9–14]. Intervening early, taking a preventative approach, and tailoring interventions to individual needs may be particularly effective [15].

The World Health Organisation has emphasised the need for evidence-based interventions to enable people with dementia to live in the community and receive tailored care [16]. However, it takes on average 17 years for an effective intervention to be integrated and implemented in practice [17] and there is a paucity in evidence relating to the implementation of evidence-based dementia care [18]. Thus, there is a need to evaluate the implementation of interventions for dementia care in practice to support rapid dissemination and scaling up of these types of intervention.

The PrAISED (Promoting Activity, Independence and Stability in Early Dementia) intervention was developed as a 12-month, home-based, individually tailored rehabilitation programme for patients with early-stage dementia or MCI, and is delivered by therapists and rehabilitation support workers, with a focus on strength, balance, physical activity and activities of daily living [15]. A feasibility trial with 60 participants showed that the intervention could be delivered as part of a research project, participants successfully recruited and followed up using research processes, and statistically significant benefits for balance and mobility outcomes [19]. A return on investment analysis showed that the intervention is likely to be good value for money [20]. A larger scale randomised controlled trial (RCT) to test the effectiveness and cost-effectiveness of the 12-month intervention, along with a comprehensive process evaluation, has been conducted [21–24].

The PrAISED research programme anticipated post-trial adoption and implementation throughout, by co-producing the intervention with health professionals, patients and their families [25–27], using health service staff rather than research staff to deliver the intervention, creating training and support resources, and by studying fidelity, adaptation and reach in a process evaluation [22, 28]. However, the challenges of the “implementation gap” are recognised along with the need to ensure the benefits for patients and healthcare services are realised widely and quickly after completion of the research. Therefore, the aim of this study was to identify what is required to implement the PrAISED intervention in a ‘real-world’ setting in routine practice in England outside of the research context. The objectives were to 1) deliver, and characterise the implementation of, a PrAISED pilot service in one National Health Service (NHS) organisation in England; 2) explore staff perceptions of the implementation of the pilot service; and 3) identify the factors which might influence the implementation of dementia care interventions like PrAISED in practice in the future.

## Methods

### Theoretical framework

The “Consolidated Framework for Implementation Research” (CFIR) [29] was used in this study. The CFIR posits that the factors affecting implementation can be related to five domains including the “intervention characteristics (innovation)”, the “inner setting”, the “outer setting”, “individual characteristics” and “process”. Across the five domains, 39 constructs have been associated with effective implementation [29]. Domain definitions and how they map to the PrAISED intervention are shown in Table 1.

**Table 1.**
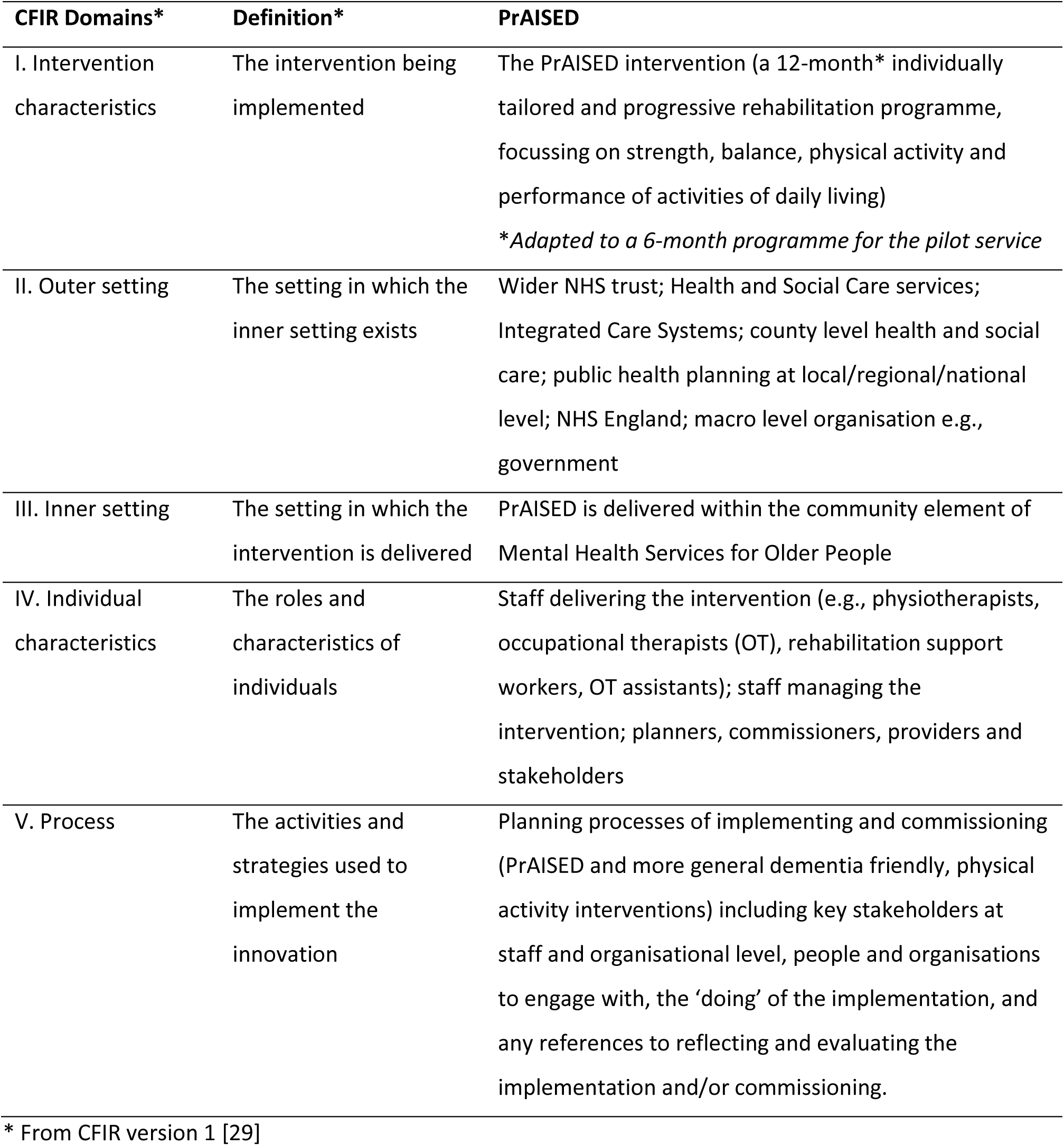
Consolidated Framework for Implementation Research (CFIR) domains mapped to PrAISED.

| CFIR Domains* | Definition* | PrAISED |
| --- | --- | --- |
| I. Intervention characteristics | The intervention being implemented | The PrAISED intervention (a 12-month* individually tailored and progressive rehabilitation programme, focussing on strength, balance, physical activity and performance of activities of daily living)<br>*\*Adapted to a 6-month programme for the pilot service* |
| II. Outer setting | The setting in which the inner setting exists | Wider NHS trust; Health and Social Care services; Integrated Care Systems; county level health and social care; public health planning at local/regional/national level; NHS England; macro level organisation e.g., government |
| III. Inner setting | The setting in which the intervention is delivered | PrAISED is delivered within the community element of Mental Health Services for Older People |
| IV. Individual characteristics | The roles and characteristics of individuals | Staff delivering the intervention (e.g., physiotherapists, occupational therapists (OT), rehabilitation support workers, OT assistants); staff managing the intervention; planners, commissioners, providers and stakeholders |
| V. Process | The activities and strategies used to implement the innovation | Planning processes of implementing and commissioning (PrAISED and more general dementia friendly, physical activity interventions) including key stakeholders at staff and organisational level, people and organisations to engage with, the 'doing' of the implementation, and any references to reflecting and evaluating the implementation and/or commissioning. |

\* From CFIR version 1 [29]

### Implementation of a pilot service

PrAISED (the intervention/innovation) was developed as a 12-month, individually tailored therapeutic programme of physical exercises, such as strength and balance training, and functional activity training [15]. In the RCT, individuals in the intervention group could receive up to 50 supervised sessions over the 12 months [21]. The current study included a 6-month version of the PrAISED intervention which was shortened due to project timelines and delivered as a pilot service.

One of the five sites which took part in the PrAISED RCT was recruited to deliver the pilot service from 1^st^ April 2022 to 30^th^ November 2022. Two physiotherapists, two occupational therapists and three rehabilitation support workers were employed part-time to deliver the service. One physiotherapist, one occupational therapist and one rehabilitation support worker were involved in both the RCT and the pilot service. The other pilot staff members were new to PrAISED and received a shortened version of the PrAISED training delivered by the research team. The therapy team were encouraged to take ownership of the intervention and make changes as necessary to deliver it as a service but were able to seek advice from the research team if required. The aim was to refer 20 patients to the service. Funding for the delivery of the pilot service was provided by Public Health England (now the Office for Health Improvement and Disparities (OHID)).

Adaptations were made to deliver the PrAISED intervention as a service. Research inclusion criteria and research processes, such as using patient information sheets and consent forms, and baseline and follow-up neuropsychological tests were removed, though the requirement for individuals to have a family member or friend to support them was retained; clinical assessments were conducted as per usual practice. A service leaflet was developed and provided to patients. The requirement for communication in English was removed as the service leaflet was made available in multiple languages and interpreters were available to support service delivery. Referrals were made to the pilot service from the NHS Trust Memory Assessment Service (MAS). The referral criteria for the service are provided in Additional File 1. The visit schedule and patient activities were adapted as the clinical team deemed necessary whilst following the principles of the PrAISED intervention.

### Study design

Quantitative and qualitative data were collected for the pilot service which included routine service delivery data; and one-to-one interviews with staff involved in delivering the intervention in the pilot service. We also conducted one-to-one interviews with staff involved in intervention delivery for the RCT at the other four PrAISED sites to gain additional insight into the perceived factors influencing implementation of the PrAISED intervention in practice.

### Data collection

#### Analysis of routine service delivery data

Routinely collected data from the pilot service were recorded on a Microsoft Excel spreadsheet by staff delivering the intervention to enable the implementation of the PrAISED pilot service to be characterised (objective 1). The overall duration of participation in the service, the total number of therapy visits and the duration of each visit were recorded for each patient. These anonymised data were sent to the research team for analysis.

#### Qualitative Interviews

Semi-structured interviews were conducted with individuals who were responsible for the management or delivery of the intervention at the pilot service site, or in the other four PrAISED RCT study sites. We interviewed managers and delivery staff at the pilot site to explore perceptions of the implementation of the service (objective 2). We interviewed staff at the pilot site and the RCT sites to gain a broader understanding of the factors which might influence the implementation of dementia care interventions like PrAISED in practice (objective 3). Four semi-structured interview topic guides (Additional File 2) were developed to reflect each type of interview (therapist/rehabilitation support worker or manager, and pilot or non-pilot site), which were guided by the CFIR (Damschroder 2009).

Clinical managers, therapists, and rehabilitation support workers at the pilot service site, and at the four other RCT sites, were invited to take part in an interview via email. Participants received an information sheet prior to the interview and gave explicit written or verbal consent. Staff from non-pilot sites were interviewed between August and September 2022; interviews with pilot site staff were conducted between October and November 2022. Interviews were conducted online using Microsoft Teams and were recorded. On average, interviews lasted 49 minutes (range 29-81 minutes). Transcripts were produced using automatic transcription and were checked for accuracy.

### Data analysis

#### Analysis of routine data

Descriptive analysis of anonymised routine data collected in the pilot service was conducted using Microsoft Excel to summarise the overall duration of participation and the mean number and duration of therapy visits.

#### Qualitative Interviews

A thematic analysis [30] was undertaken using a pre-determined CFIR codebook to guide the analysis [31]. The analysis was conducted in NVIVO [32]. Transcripts were read and re-read by two researchers (CB & JL) to aid familiarisation with the data. Initial coding was conducted by applying the CFIR codes to each transcript. Notes were made on the transcripts to justify coding decisions. EA checked coding throughout to facilitate a consistent application of CFIR. Regular weekly meetings were held with the wider team (CB, JL, EA, RV, RT) to discuss coding and emerging insights. Potential themes and subthemes were discussed and revised iteratively to produce a final set of themes relating to each of the CFIR domains. During our study, an updated version of the CFIR was published [33]. We included new or revised construct definitions where these were deemed relevant to our study.

## Results

### Pilot PrAISED service (Objectives 1 and 2)

Eleven patients were referred to the service. Staff delivered 230 visits in total (mean 21 visits per participant, range 5-39) with a mean visit duration of 82 mins (mean range 68 to 91 minutes). The overall dose of intervention varied depending on when the participant was recruited: eight participants received six months intervention, one participant received five months, and two participants received four months intervention.

Seven interviews were conducted with staff from the pilot service (two managers, two physiotherapists, two occupational therapists and one rehabilitation support worker). Five themes were identified across seven CFIR constructs (Table 2): operational processes; workforce capacity; referral; intervention delivery; and patient impact.

**Table 2.** CFIR Domains and constructs associated with themes in the Pilot Servic.

| Theme | CFIR Domain* | CFIR Constructs* | Key points |
| --- | --- | --- | --- |
| Operational processes | III. Inner setting | A3. Work infrastructure | Operational processes took much longer to establish in the pilot service |
| Workforce capacity | III. Inner setting | E2. Available resources | Small, part-time team, working on PrAISED alongside normal clinical role, made it challenging to meet the needs of the service |
| Referral | V. Process | B2. Innovation recipients | There were challenges with referrals to the service<br>Influenced by the short time frame for the pilot service<br>Potential difficulties engaging the referral team / challenges with duration of appointments to cover all services available to patients<br>The timing of referrals may have impacted patient engagement; some patients needed more time to come to terms with their dementia diagnosis and may have started PrAISED at a later date |
| Intervention delivery | I. Intervention characteristics | D. Adaptability | 6-month pilot service was more like normal practice, with a shorter intervention and more flexibility for patients to join and leave the service<br>There was flexibility in delivering the service, whilst retaining the PrAISED intervention principles<br>It was challenging to fit in the visits in the shorter time frame<br>Time constraints made it difficult to build rapport with patients; and led to a change in focus of activities/visits<br>Less focus on rehabilitation and more on risk assessment/management, quick strategies, and physical elements/exercises<br>Goal setting continued, but these had to be prioritised; it was harder for patients to achieve goals in the shorter time frame<br>Less use of dual tasking elements / used more towards the end of the service<br>Lack of time to taper visits, more reliance on written materials and signposting to other services<br>Dependence on referral to other services for sustainability |
|  | V. Process | C. Executing/ doing |  |
| Patient impact | II. Outer setting | A. Needs & resources of those served by the organisation | The service was well received by patients, with most providing positive feedback as to perceived improvements in confidence, motivation, balance and strength, and reduced loneliness |
|  | V. Processes | D2. Reflecting and evaluating – intervention |  |

\* From CFIR version 1 [29]

#### Operational processes

Findings from the interviews suggested that establishing operational processes for the pilot service was more complicated and took longer than in the RCT requiring buy-in and approvals from the wider organisation.

> *“All the operational things that you’re relying on Trust colleagues to get back and confirm, rather than when we worked on the RCT when it was between the research team, the CRN [Clinical Research Network]. It’s just those kinds of procedural things just took a bit more thinking and organising I suppose.”* (Manager)

#### Workforce capacity

Scheduling multiple therapy sessions at a time acceptable to patients, and communicating between a multi-professional team, in the face of competing demands on time and part-time working, was challenging. However, recruiting lower numbers of participants than expected made it more manageable for the small team employed to deliver the service.

> *“Everybody’s trying to do it as part and parcel of something else that we’re doing, so we had to work quite hard at staying in touch with each other. So that’s been a bit of a challenge.”* (Occupational Therapist)

#### Referral

The pilot service did not recruit its target number of participants (11 patients were recruited from the target of 20). Referral to the service was thought to be challenging due to the nature of the pilot service (short-term and temporary, so there was no time or incentive to embed processes in clinical activities); high turnover in the MAS referral team; and keeping referral staff up to date with the rapidly changing services available.

> *“I was surprised we didn’t get more referrals. But I think it’s interesting to hear from some of the other studies that’s quite normal, and it’s a reflection of staff on the ground, those referring in, to get their head around a new offer or a new service.”* (Manager)

In addition, the referral team had time constraints during appointments with newly diagnosed patients, and there was a large amount of information about services and research studies they could potentially cover alongside conducting assessments and providing a diagnosis. This was thought to make it difficult to always include PrAISED in the discussions.

> *“Some of the people that recruited have worked in the MAS service and were very proactive, really understood the study and were really effective about recruiting, whereas other staff, I think they get so many studies and requests for a memory assessment, and also the time that they’ve got to interview people and talk about everything is really short.”* (Manager)

Furthermore, the volume of information for patients to take in at the point of diagnosis was also thought to be a challenge and might have influenced patient’s decisions as to whether to take part in the PrAISED service.

> *“They’ll [the patients] have come home with ‘What is Alzheimer’s disease? What is this? What is …, you know? If I need some advice about my …, I’ve got all these bits of paper. I’ve just been given a diagnosis.’ It’s almost too much to take on at that point in time.”* (Manager)

#### Intervention delivery

Interviewees felt the pilot service was more like normal practice in duration, but there was more flexibility than in the RCT in the number, duration and frequency of visits with participants.

> *“I think every visit is very unique for every person. Sometimes a visit can take 40 minutes, sometimes if the individual is struggling, you do tend to give them a little bit more time. So, I think having that flexibility has been incredible because there’s no set time of going between visits.”* (Physiotherapist)

In comparison to the RCT, there was also more flexibility in timing for participants to join the service or to leave the service if there was a lack of engagement or motivation or the service wasn’t thought to be right for the patient.

> *“We said, ‘Okay well, now this isn’t useful for you’ so we’ve signposted to where we felt was better for that person. We felt like she needed a care package in the end, and that was more what her needs were.”* (Rehabilitation Support Worker)

Staff who had worked on the 12-month intervention in the RCT reported finding the shortened duration of the 6-month pilot service intervention challenging. It impacted on fitting the visits in and delivering such an intensive intervention, though staff reported they continued to follow the principles of PrAISED in delivering intervention activities.

> *“Everything has been halved in terms of the usual number of contacts that we were able to have within a year. I know we’ve struggled to get the amount of RSW [Rehabilitation Support Worker] visits in. There’s been no change to the focus of the interventions that we’ve been we’ve been doing, but we’ve had some time constraints in terms of being able to do things as thoroughly as we were able to do when we had a whole year to work with people.”* (Occupational Therapist)

The shorter duration of the pilot service intervention was thought to change the focus of visits in some cases. There was less time to build rapport with patients, complete assessments and set goals, but staff used their skills and experience to innovate and adapt delivery of the intervention. There was less focus on rehabilitation activities and more risk assessment/management, providing written materials, quick strategies such as physical elements/exercises and signposting to other services to provide long-term support.

> *“It takes time to build up rapport with somebody, to do your assessments, to plan goals, to see what things they value, and I think that’s been much quicker, and I think there’s been less actual rehabilitation and a little bit more signposting earlier on, when they haven’t had the benefit of the actual rehabilitation intervention.”* (Manager)

Interviewees reported that goal setting continued, but these had to be prioritised due to the shorter time frame, and it was harder to achieve goals in the time available.

> *“I think with the… study [RCT], because we could work with people for a year, one of the key things that is really important is being able to negotiate and set goals with participants and ask ‘What do you want to achieve?’ and usually the participant will say, ‘Well, I want to achieve this within a year’. With this [the pilot service], we’re trying to help somebody set goals that are a bit shorter because you’ve only got three to six months to work with somebody.”* (Occupational Therapist)

One of the key elements of PrAISED was to build in dual tasking (activities which involve simultaneously performing two or more tasks, such as a motor task and a cognitive task) and apply this to the exercise programme adapting it to the needs and abilities of the patient. There was less use of dual tasking activities in the pilot service, or these were used more towards the end of the intervention.

> *“Mostly towards the end of the contact time with somebody, I’ve been able to start building in dual tasking, because we’ve been working on other things that have been more of a priority. So, the dual tasking element probably slipped a little bit in terms of when we’ve been able to bring that into the programme.”* (Occupational Therapist)

Staff delivering the interventions recognised the importance of the tapering and ensuring the patients did not become dependent on them, however they found it much more difficult to do this within the shorter time frame of the pilot service.

> *“I think the tailing off part is really important because you do develop that relationship with the individual and because you are there supporting them through a time which is very difficult, and I can imagine quite scary as well for them. So, that tailing off period is very important.”* (Physiotherapist)

Due to time constraints in the pilot service, there was more dependence on signposting, provision of materials and referral to other services to support activities and delivery of the intervention, and for sustainability beyond the end of the service.

> *“We’ve been trying to make sure we’re leaving people with as many resources, psychologically and practically, to be able to carry on when we’re not there. So, we’ve tried very much to link them in with other community resources and community services that are going on that they can access.”* (Occupational Therapist)

#### Patient impact

Staff perceived there were benefits to participants from the intervention. They reported the service was well received by patients, and it made them feel valued with most providing positive feedback and reporting to therapists perceived improvements in confidence, motivation, balance and strength and reductions in loneliness.

### Factors influencing the implementation of PrAISED in practice (Objective 3)

A further eight interviews were conducted with staff from across the other four RCT (non-pilot) sites (two per site, one manager and one member of intervention delivery staff) which were combined with the interviews with staff who delivered the pilot service to identify themes related to the wider implementation of PrAISED and similar interventions. Nine participants were therapists or rehabilitation support workers and six were managers. From these fifteen interviews, six themes relating to implementation issues were identified across twenty-two CFIR domains (Table 3): the need for support post-dementia diagnosis; acceptability; effective delivery; reach/referral; intervention design and adaptability; and intervention materials and training.

**Table 3.**
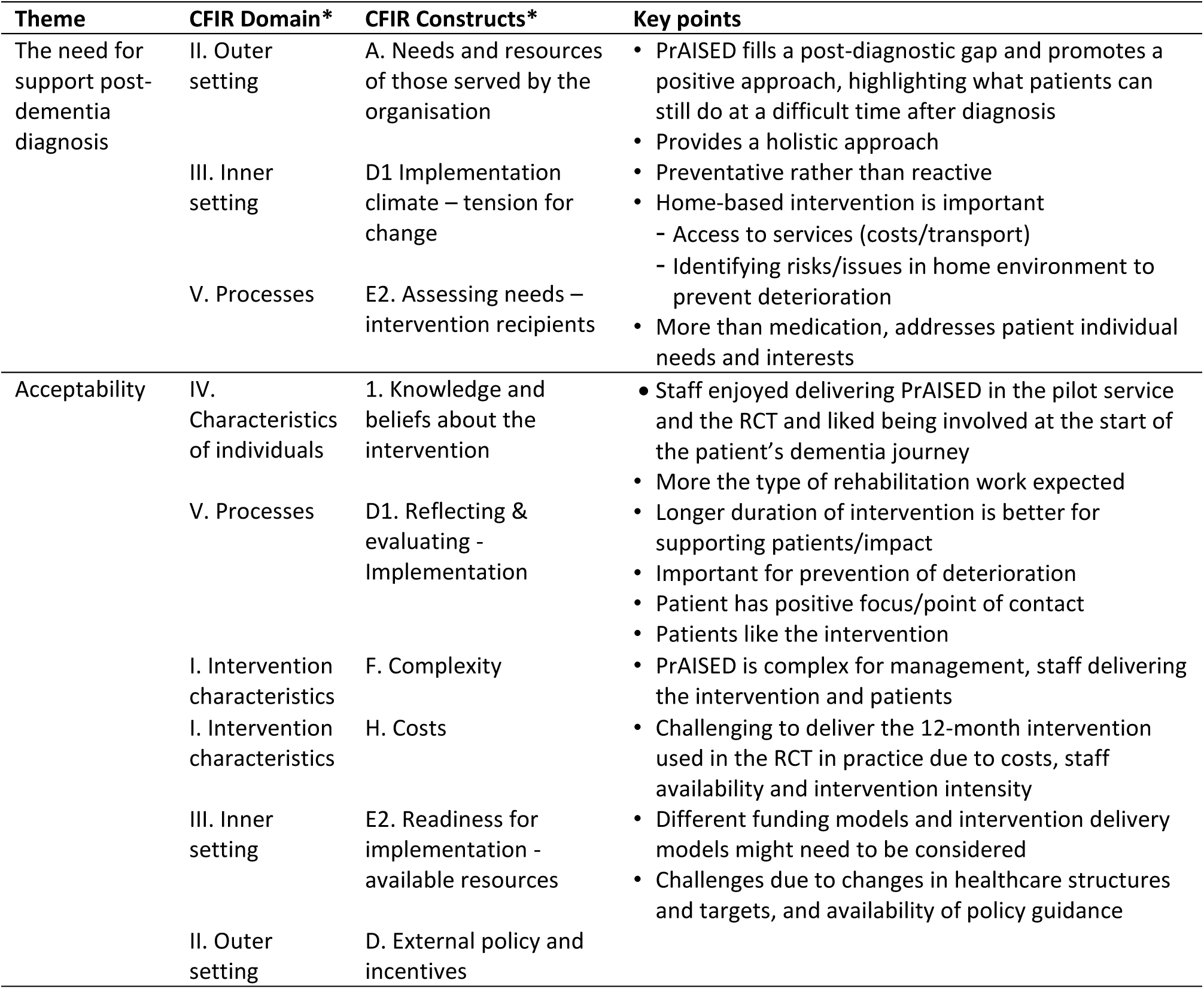
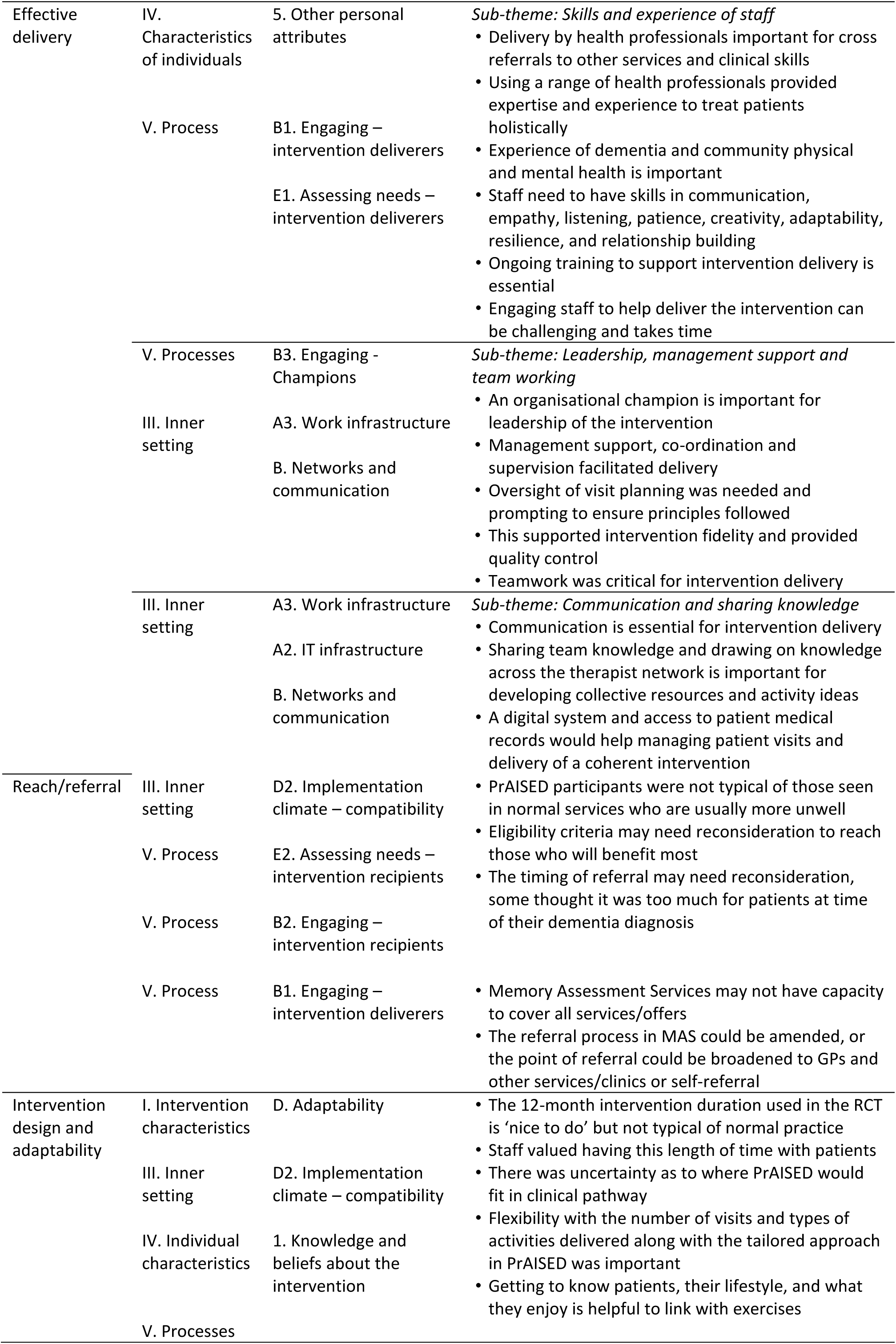
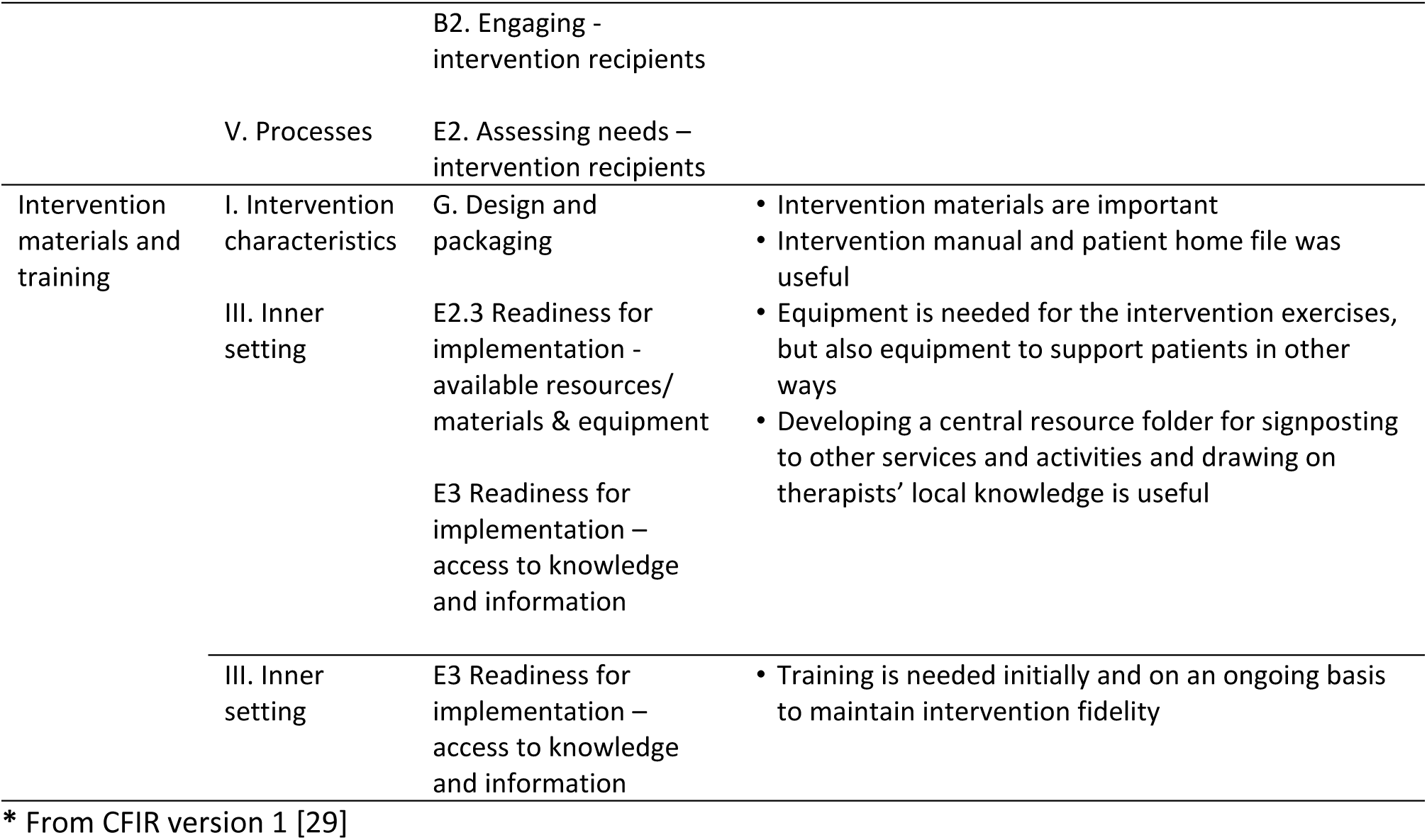
CFIR Domains and constructs associated with themes for PrAISED implementation.

#### The need for support post-dementia diagnosis

Interviewees thought PrAISED fills a gap immediately post-diagnosis, and promotes a positive approach, highlighting what patients can still do and working towards bigger goals at a difficult time after receiving a diagnosis. It provides a tailored, holistic and preventative approach for patients with dementia, intervening at an early stage in a patient’s diagnosis. This contrasts normal services which are typically more reactive and get involved when a patient reaches crisis point.

> *“It’s just so evident there’s a gaping hole, and it is at the most valuable time to engage with people and to have that dialogue with individuals when they have capacity to do so, because often by the time CMHT [Community Mental Health Team] get involved, people will have limited capacity and difficulties consenting to decisions.”* (Occupational Therapist)

The home-based nature of PrAISED was thought to be important in reducing issues related to accessing services. For example, some patients can’t travel to venues for sessions due to costs and availability of transport, potentially widening health inequalities. Interviewees also felt the home-based nature of the PrAISED intervention was important because it enabled them to identify risks or issues for patients in their home environment that might not be observed in a clinical setting, or until a later stage when the patient had deteriorated.

> *“This programme is all about really seeing somebody in their home environment and looking at the risks and looking at their challenges and sometimes you can notice things around the home that you can build into the programme.”* (Occupational Therapist)

Intervention delivery staff indicated they want to be able to offer patients more than medication, and to have the time and flexibility to work with patients, tailoring the number of visits and the activities chosen to meet the needs and interests of the patient with dementia to enable them to participate, and motivate them to do the activities. PrAISED was thought to provide this opportunity.

#### Acceptability of PrAISED

Staff enjoyed delivering PrAISED therapy and thought it was more the type of rehabilitation work they expected to do when they trained as therapists. They liked being involved at the start of a patient’s dementia journey and seeing the impact it had.

> *“It’s really a privilege to do as a therapist because you’ve got time to really get to know the person and explore their individualised needs. You’ve got time to adapt it and change it, and you know that’s not something I often get in any other role of therapy. It’s lovely from a therapy delivery point of view, but also, it’s lovely seeing the difference that makes to people.”* (Physiotherapist)

Interviewees believed that early intervention immediately post-diagnosis and a sufficiently long duration of intervention was important for supporting participants, helping them achieve their goals, preventing deterioration and increasing the intervention impact, giving participants a positive focus and a point of contact. There was a perception that patients also liked the intervention.

> *“What I’m hearing from the individuals and from their carers is that PrAISED provides hope and it’s a real positive part of their journey because we’re looking at positive things with them. We’re concentrating on what they can do, not what they can’t do, and we’re concentrating on where they want to go, and they can see that there’s a progression in a positive way that it isn’t just about deterioration. But what PrAISED is really good about, is highlighting what are the small achievable goals and even bigger goals that you can continue to strive towards, and that there are ways around problems. It seems just the perfect time really to capture those people.”* (Occupational Therapist)

In contrast, there was also some perception that the PrAISED intervention is complex which may impact on future implementation. This was due to the referral process, the number of different staff involved, the number and varying nature of patient visits, the management input required to ensure visits are being undertaken, and making sure patients adhere to the intervention. In addition, it was thought to be complex for patients due to interchangeable use of terminology, e.g., goals/aims or exercise/activities, and setting goals and using dual tasking; and this might need to be simplified in future.

It was thought it would be challenging to deliver a 12-month intervention in practice due to costs, staff availability, and the intensity of the intervention (up to 50 visits), and interviewees were uncertain how this could be funded. Interviewees thought different funding models and different ways of delivering the intervention would need to be considered. For example, how to train staff; how to maximise the use of different types of staff (e.g., occupational therapists, physiotherapists and rehabilitation support workers) with different skills cost effectively; how to reduce the costs of travel to patient’s homes (whether some of the intervention could be delivered remotely); and whether a more flexible budget could be provided that could be used as needed for patient resources and equipment. It was suggested that some investment would be needed initially to support developing and establishing the intervention as a service.

> *“I think internally it will be very difficult until things got established. It needs pump priming because I think some of our resources could be used in a different way. But until you invest in it, you can’t release those resources. It needs some sort of investment to start and to get some committed staff time to deliver the intervention, as it’s supposed to be delivered and not a watered down version.”* (Manager)

Furthermore, it was thought that recent changes in the health service structure in England, particularly in the way services are commissioned, along with regular changes in initiatives and targets, the need for evidence of effectiveness and cost-effectiveness, and uncertainties of where dementia services sit within the NHS may make it challenging to embed PrAISED into practice at present.

#### Effective delivery

Three key areas were identified in relation to effective delivery including the skills and experience of staff; management support and team working; and communication and sharing knowledge.

##### Skills and experience of staff

Interviewees thought PrAISED should be delivered by health professionals. They have the clinical skills needed to work with patients with dementia so can address other medical issues and have awareness of patient safety and knowledge of additional services where patients can be referred to if other types of support are needed. Having a range of health professionals with different areas of expertise was thought to be important as it enabled the team to have wide-ranging and comprehensive knowledge and experience to ensure patients were treated holistically.

It was thought that staff delivering the intervention should have experience of dementia and of community physical and mental health, as they complement each other and enable more comprehensive approach to treatment. Having skills in communication, empathy, listening, patience, creativity, adaptability, resilience, and relationship building were thought to be essential, along with ongoing training and shadowing/supervision of sessions for staff delivering the intervention.

> *“It would be useful to have some experience of working with dementia and possibly useful to have some experience working in the community as well, and those people skills, listening and trying to understand, being empathetic.”* (Physiotherapist)

Furthermore, interviewees reported that engaging staff to help deliver the intervention can take considerable time. Having good links with the correct services and providing reassurance to potential intervention delivery staff regarding workload, protected time, support and training is needed. Having sufficient staff capacity to deliver the intervention to many patients simultaneously was thought to be one of the main barriers for delivery.

> *“It takes a lot of time to engage clinicians, letting them know that they have got us for support and that it’s all about their protected time. There was lots of reassurance.”* (Manager)

##### Leadership, management support and team working

Leadership, management support, co-ordination and supervision along with team working and regular communication was a facilitator in delivering PrAISED. Having a champion within the organisation who could lead and promote the intervention, obtain approvals and ensure the quality of the intervention delivery was critical.

> “*You need to find one good person to take up your cause within the department, that has good relationships with everybody and can see the benefit of the intervention. You really need that one champion initially, who can bring people on board and help you.”* (Therapist)

It was important for management to oversee visit planning to ensure visits had been completed in the correct timeframes, and to prompt staff regarding use of the intervention manual to ensure the PrAISED principles were followed to support intervention fidelity and provide additional support for intervention quality control.

> *“It needs management. It needs somebody supervising that the visits are being taken as per the schedule and that they are time effective as well.”* (Occupational Therapist)

A collaborative relationship between the occupational therapists, physiotherapists and rehabilitation support workers, who jointly delivered the intervention, was important for planning visits, delivering sessions and monitoring patients’ progress.

##### Communication and sharing knowledge

Communication was essential for effective delivery of the intervention. This included team meetings to review cases, keeping an oversight of patient management and sharing strategies and ideas for activities; sharing knowledge and information about services and activities outside of PrAISED; communication to patients about their participation; and communication to general practitioners (family doctors) at the end of the intervention to share progress and a future action plan.

Using team knowledge, sharing ideas and drawing on knowledge across the therapist network (particularly if a therapist was working in a geographical area that was not familiar to them) was important for developing collective resources and pooling information about ideas for community activities for patients, along with services which were available where patients could be referred or signposted to support delivery of PrAISED and long-term participation beyond the end of the intervention.

> *“I think it worked well because the rehab worker lived in the area, she knew we don’t live in the area, so we didn’t know what was available, but she was brilliant, she knew loads so we were able to tap into her local knowledge.”* (Occupational Therapist)

It was also suggested that digital systems would be useful for managing patient visits along with having access to patients’ medical records, which was considered important to facilitate delivery of a coherent intervention across different members of the team and in collaboration with other services being offered to the patient.

> *“With [County1], we had no access to the electronic patient record system, which meant that for most of the referrals, we were working blind, we weren’t able to read up on any previous medical history or previous mental health history, or even ongoing contact with services. In [County2], we’ve used [electronic patient record system] all the time and I have found that very helpful. I think it’s really important that any new service has to be integrated, so staff can see the patient record.”* (Occupational Therapist)

#### Reach/Referral

Interviewees thought patients in PrAISED were not the typical group of patients that would be seen and that usually patients would be seen when their disease had progressed more and were more unwell.

> *“They wouldn’t meet the inclusion criteria; they’d be too high level and they’d be able to get to other services. So, it’s a real privilege to be seeing people that are able to do such high level and quality of life intervention, but it wouldn’t fit with the other service that I work in currently, we would not have the capacity to see people that were that able.”* (Physiotherapist)

It was suggested the eligibility could focus on need more, for example people who are less physically active, to reach those who will benefit most, along with some form of motivation assessment to ensure patients engage in the intervention.

> *“When we were doing PrAISED, we did get a number of people who were already quite active that didn’t necessarily need as much intervention as they were already going to the gym four times a week. You’re not going to refer somebody for a service that they don’t need, are you? So, you would be checking their activity levels before the referral was made.”* (Manager)

There were mixed views about the timing of referrals to PrAISED. Some thought it was best to refer at the time of the diagnosis appointment when patients most needed support, whereas others thought it may have been too much for patients at this point. It was suggested that the referrals process is not currently designed with the patients’ needs in mind and needs to be later in the pathway as the patient can be overwhelmed at their point of diagnosis.

There were also mixed views about the process of referrals to PrAISED. There were concerns that Memory Assessment Services (MAS) may not have capacity which may reduce referrals to the service. As noted in the pilot service, MAS had lots of things to cover at the appointments already and there was little time to mention additional services in detail. This along with high staff turnover means that regular training and reminding of the services that are available are thought to be needed. Several suggestions were made for alternative referral approaches. These included introducing separate appointments at the MAS, one for diagnosis, and one for next steps/support (either face to face or by telephone); having a PrAISED representative (or someone to represent PrAISED e.g., from the Alzheimer’s Society) in clinics for patients to speak to after their appointment; a follow-up telephone call; and broadening referral points to include GPs and other services, for example a social prescribing service, other clinics, e.g., memory clinics, a falls clinic, or day centres, or alternatively, self-referral to the service.

> *“But that’s what the Alzheimer’s Society could do. It doesn’t need to be a psychiatric nurse or an OT, or a physio explaining the service offers, and actually, to have someone in contact with the Alzheimer Society at that point in time, I think it’s really good because they are accessible.”* (Manager)

> *“It would be interesting if it was a self-referral, so if you went to a memory clinic, a memory cafe for example, and there were lots of leaflets about where people could refer to, and some people can self-refer themselves to NHS services”* (Occupational Therapist)

#### Intervention design

Staff liked the original 12-month duration of the intervention delivered in the RCT and valued this time with patients but thought this was ‘nice to do’ and not typical of normal practice. They would normally have a much shorter timeframe to work with patients, e.g., 6 weeks, and the patients would be discharged as soon as they had completed the short intervention to make space for the next patient.

> *“You were given a client for six weeks and in that six weeks you set goals and implemented them; and if they didn’t complete it, you would signpost them to different ones.”* (Occupational Therapist)

Interviewees were unsure where PrAISED would fit in normal clinical pathways and there were mixed views as to whether it should be integrated into existing services or kept as a separate service, with some being concerned ‘it would get lost’ if it was integrated. The flexibility and tailored approach to visits and activities in PrAISED is important and it was seen to be positive that the intervention was not limited to a fixed number of sessions in a fixed number of weeks followed by discharge from the service because there are other patients on the waiting list. Getting to know patients, their lifestyle and what they enjoy is helpful, including linking back to things the patients used to do, as these can be linked to the intervention activities.

#### Intervention materials and training

Having resources to support the delivery of the intervention was very helpful. This included an intervention manual, patient home file and equipment, both for the intervention exercises and to support participants in other ways e.g., rails, plastic step by shower. The intervention manual was thought to be very comprehensive and was a ‘bible and reference point’ for the intervention. Staff used it in different ways, some ‘dipping in and out’, but they thought it was important for ensuring the intervention was delivered as prescribed and not just ‘ad hoc’. The intervention manual was thought to be invaluable for supporting training.

The patient home folder (an A4 size ring binder folder) was thought to be ‘brilliant’, and most patients found it useful and took pride in having it. Interviewees reported that the file was however quite big, and some of the paperwork and terminology was confusing for patients. It was suggested that a smaller, simplified exercise booklet might be helpful where staff delivering the intervention can tick off the exercises to do. Interviewees thought the folder was useful for giving patients information to use in the future, even if it was not used during the intervention. Some staff developed a central resources folder to support signposting to other activities and services during the intervention, though this took time to build up and required local knowledge and experience. Other interviewees used a search engine such as Google to find services. Interviewees thought it would be useful to set up a central folder or database in future for signposting/what services are available in each local area. Sharing knowledge and making use of therapist networks was useful to support this wider knowledge of community resources and how to access these. It was also noted that sometimes there isn’t a service that meets the patient’s needs in their area.

> *“There’s so many things out there, so many charities, so many resources. I think there’s a central resource hub now where you can have a look. So, if you’re looking for a dementia swimming group for example, you can just search yourself.”* (Occupational Therapist)

Whilst most staff already had experience and skills in working with patients with dementia, specific PrAISED training was provided for the research study and the pilot service by the research team. Staff delivering the intervention received different formats and amounts of training depending on when they joined the study. Some received three days face to face, whereas others received two days remotely (mostly due to the COVID-19 pandemic social distancing restrictions); most received refresher training which was also thought to be useful. The training provided was reported to be very good quality and well delivered. Interviewees reported preferring face to face training as it allowed more networking and sharing than remote training. Others liked the online training as it allowed time to pause the session and write notes. Shadowing colleagues was also thought to be useful to increase confidence with delivering the sessions.

> *“Having a bit of training is definitely important, the training was really helpful for me initially to understand what the aim and what the goals were, what available advice there was out there and how to tailor my visits, and what to do on each visit was really helpful.”* (Physiotherapist)

Interviewees thought that future training should be offered online and face to face but there was uncertainty as to who could provide this in practice. It was felt that core training was needed but local training may be required to address operational differences in different organisations, and ongoing training should be offered to ensure fidelity of the intervention. It was recognised that staff have differing backgrounds, and that training may need to be tailored to take this into account. It was also thought that shadowing an experienced therapist on visits/assessments when new to the intervention to see processes and understand how to deliver the sessions was important.

## Discussion

There is a paucity of evidence relating to the implementation of dementia care interventions [18]. PrAISED was developed as a 12-month, home-based, individually tailored rehabilitation programme for patients with early-stage dementia or MCI, and is delivered by therapists and rehabilitation support workers, with a focus on strength, balance, physical activity and activities of daily living [15]. The aim of the current study was to identify what is required to implement the PrAISED intervention in routine practice to develop the evidence base for the implementation of dementia care interventions. We explored this through delivering a 6-month version of the PrAISED intervention as a pilot service and conducting interviews with those delivering PrAISED as a service, as well as interviewing those involved in delivering the intervention provided during an RCT to gain wider insight into the factors that might influence implementation in routine clinical practice.

We found it was feasible to deliver a 6-month PrAISED intervention as a clinical service with some adaptations, including removal of the research inclusion criteria and replacing them with referral criteria, and removal of research processes (e.g., replacing the patient information sheet with a service leaflet) to make it suitable for service delivery. However, referrals were fewer than expected and there were some changes to the focus of the intervention activities. The possible reasons for this are discussed below and require further investigation.

From qualitative interviews with staff involved in both the delivery of the PrAISED pilot service and the RCT intervention, we found themes which align to the CFIR and show implications for the future implementation of the PrAISED intervention, and other similar dementia care interventions, in routine clinical practice. These include the need for: patient support and preventative interventions immediately post-dementia diagnosis; funding to establish and implement interventions; leadership and management, including ‘champions’ who can promote dementia care across organisations and the wider healthcare system; time to establish operational processes and embed the intervention into organisational pathways; time to engage relevant staff with wide-ranging personal and clinical skills, provide staff training and establish collaborative working relationships; development of intervention resources and processes (including access to patient medical records), and identification of local services and activities which can support intervention delivery; referral criteria to be established along with referral procedures for suitable patients, and referral staff to be trained on an ongoing basis; and, ensuring interventions can be adapted and delivered flexibly to meet the needs of delivery staff and patients whilst maintaining fidelity to the original intervention where possible. These are discussed in more detail below.

### Support post-dementia diagnosis

Our study confirms there is a gap in support for patients immediately post-dementia diagnosis. This has been reported previously [34–37] and exists despite the policy focus on dementia [38]. Whilst the gap may partially be due to lack of infrastructure, and capacity and capability in services [37], should these infrastructure and resource issues be resolved, PrAISED or a similar intervention could fill this need for support by intervening early and providing a holistic and preventative approach for promoting physical and mental health in dementia patients.

### Funding, leadership and operational processes

In order to establish and deliver PrAISED or similar dementia care interventions in current UK healthcare systems, funding will be needed with a substantial initial investment to establish resources and staffing for the intervention. There is uncertainty as to how interventions like this could be funded and integrated into current service provision, which will be a major barrier to intervention implementation and scale-up. The challenges with commissioning preventative interventions such as PrAISED have been reported elsewhere [39] with similar findings identified in another UK-based intervention designed to support patients and their carers post-dementia diagnosis [40].

A ‘champion’ is needed to promote dementia care interventions and provide leadership and management across organisations and the wider healthcare system. Evidence suggests there is an association between the use of champions and increased use of healthcare innovations by organisations [41], and that leadership and management support facilitates implementation of dementia care [18]. To facilitate the effective implementation of any new dementia care intervention, and before commencing service delivery, time is needed to establish operational processes (which may take longer than the processes which need to be put in place for research studies), to identify how best to embed the intervention into organisational pathways, to engage relevant personnel and intervention staff (for referral and delivery) and establish working relationships, and to provide staff training. Leadership, the time and effort needed for preparation for implementation and adoption of new interventions, and organisational readiness for change are critical elements of implementation success [42–44].

### Staff skills, intervention training and resources

Findings from this study suggest that staff delivering PrAISED or similar dementia care interventions should be healthcare professionals with experience of working with patients with dementia along with wide-ranging personal and clinical skills. This contrasts with the views of commissioners and other key stakeholders’ who thought that non-health professionals, such as exercise professionals, could also potentially deliver the intervention to increase the workforce capacity [39]. Further investigation is required to determine if these other professionals, e.g., those working in the sport and exercise industry, could play a role in delivering PrAISED or similar interventions.

Staff need to have capacity to deliver the intervention and be able to work collaboratively as part of a team. Teamwork is essential for effective intervention delivery, and this is known to be important for providing patient care [45]. Intervention specific training is needed for staff referring to any new service as well as those delivering the service; it is not clear who would provide this training, and this requires further exploration. It has been noted that the time and support needed for staff to gain experience and confidence in delivering a new intervention should not be underestimated [46]. Resources such as training materials, an intervention manual, a patient file, and equipment are important for intervention delivery, along with having access to patients’ clinical records, so those delivering the intervention are aware of patients’ medical history, their wider needs and any ongoing support being provided by other services. In addition, therapist knowledge is important for identifying local services and activities to support intervention delivery and sustainability of dementia care.

### Patient eligibility and referral

The timing of referral to the PrAISED intervention at the point of diagnosis meant that some of the patients referred to PrAISED were still physically very well and able. Typically, they would not be seen in normal services until their disease had progressed, they had deteriorated and required assistance. However, as noted above, there is a clear need for interventions immediately post-diagnosis as a prevention strategy and to support patients at this time, regardless of their physical status. Commissioning preventative interventions, such as PrAISED, for patients with dementia at the time of their diagnosis requires a shift in focus for healthcare services to facilitate access to funding and delivery of interventions of this type. Whilst the NHS Long Term Plan [47] highlights the contribution the NHS will make to prevention activities, it may take some time for this healthcare transformation to be realised.

There were challenges in receiving referrals to the PrAISED pilot service, and this has been noted in other similar studies [46]. Referral pathways need further investigation to increase the number and diversity of patients referred to dementia care services like PrAISED. Ethnic and socio-economic diversity in PrAISED was limited, and research has shown there are substantial barriers to engaging in dementia healthcare, rehabilitation, and research for some ethnic groups (Howe et al.; forthcoming). Options for increasing referrals might include considering whether staff in MAS have capacity to undertake referrals to other services alongside their other activities and how this could be managed, and broadening out points of referral, for example to GPs, memory clinics, falls clinics, day centres or self-referral. The timing of referrals to best support patient needs also needs to be considered. Offering flexibility in the timing of when a patient starts a dementia care intervention may help to address patient preferences for when they most need the support.

### Intervention design and fidelity

Home-based dementia care interventions such as PrAISED may help to address inequalities in access to healthcare services for some population groups, as well as facilitating the assessment of patients with dementia and supporting them in their everyday environment. The shortened duration of the pilot PrAISED intervention (due to project timelines rather than a desire to change the 12-month duration of the intervention tested in the RCT) impacted on intervention delivery. There was increased flexibility with regards to intervention delivery, but there was a reduced number of visits and there were some changes in session content due to having less time and fewer weeks to work with patients. The changes included more focus on risk management, provision of written materials and signposting or referral to other activities and services, prioritising goals, and there was reduced time for tapering therapists’ involvement. The duration of any intervention offered in clinical practice would ideally be long enough to allow time to effectively work with patients and enable them to achieve their goals, and deliver the intervention activities as planned, with flexibility for delivering visits and adapting the intervention to patient needs. Balancing intervention adaptation with intervention fidelity remains a challenge in health promotion and prevention but using adaptable designs, rather than designs with strictly defined fidelity criteria, are thought to be more sustainable and more likely to have public-health impact [48].

### Strengths and limitations

This study included a ‘first attempt’ at delivering the PrAISED intervention as a service in practice. It adds to a paucity of data regarding the implementation of interventions for patients with dementia and provides insight as to what may be required to deliver an evidence-based dementia rehabilitation intervention in routine clinical practice. The pilot service was delivered by an organisation which was involved in the RCT evaluating PrAISED and therefore they may have already been bought in to the intervention, have staff and systems in place and be familiar with intervention delivery. Whilst this facilitated intervention implementation in the pilot service, there will be organisational barriers to delivering PrAISED in organisations who are completely new to the intervention. The length of the pilot intervention was constrained by the end date of the PrAISED study, which may have affected the number of referrals to the service and fidelity. Many interviewees discussed the difficulties of delivering this shorter intervention to patients; however, this may be more reflective of what could be feasibly funded and delivered in NHS services. Given the short duration of the study, the available funding, and the focus on implementation of the intervention, we did not assess the impact of the intervention on patient cognitive and physical outcomes. A future hybrid effectiveness-implementation study is needed to explore this further. Staff who may have been less enthusiastic or motivated to deliver the PrAISED intervention may have been less likely to participate in the interviews.

## Conclusion

There is a need for interventions to maintain physical and mental health, and social engagement immediately post-dementia diagnosis. It was possible to deliver a 6-month version of PrAISED as a service in practice, but adaptations were required to deliver the intervention as a service instead of a research study; and referrals were lower than expected. Future implementation of PrAISED or similar dementia care interventions will require attention to identifying intervention funding, leadership and management, time to establish operational processes, the skills and experience of intervention deliverers, providing training and resources to support intervention delivery, patient eligibility and referral processes, and the duration and components of the intervention. Future research might include a hybrid effectiveness-implementation study to explore these issues further, to examine intervention adaptation and fidelity in practice and to assess the impact on patient outcomes to increase the evidence base for the implementation and effectiveness of dementia care interventions in practice.

## Data Availability

All data produced in the present study are available upon reasonable request to the authors.

## List of abbreviations

CFIR: Consolidated Framework for Implementation Research
MAS: Memory Assessment Service
MCI: Mild Cognitive Impairment
NHS: National Health Service
PrAISED: Promoting Activity, Independence and Stability in Early Dementia
RCT: Randomised Controlled Trial
UK: United Kingdom

## Declarations

## Ethics approval and consent to participate

The current study received research governance approvals and ethical approval from the Bradford Leeds Research Ethics Committee as a sub-study to the main PrAISED2 trial (18/YH/0059; 236099) on 25^th^ May 2022. Approval was also obtained from the NHS Trust participating in the pilot service for service delivery and analysis of data relating to intervention delivery. All interviewees gave written or verbal consent to participate.

## Consent for publication

Not applicable.

## Availability of data and materials

The datasets analysed during the current study are available from the corresponding author on reasonable request.

## Authors’ contributions

EA, RV, ST, EO, SG, JG, TM and RH designed the protocol for the implementation study. HS led the implementation of the pilot service and oversaw recording of data for the service. EA analysed the quantitative data from the pilot service. CB and JL conducted the interviews. EA, CB and JL coded and analysed the interviews and identified recurring themes. EA, CB, JL, RV and RT discussed the findings to facilitate data interpretation. EA drafted the manuscript. All authors critically reviewed and edited the manuscript and read and approved the final version.

## Acknowledgements

The authors wish to thank the staff and patient participants in the PrAISED implementation study for their contribution to the project.

## Additional Files

### Additional File 1

Filename: Adams Additional File 1.pdf

Title: Referral criteria for PrAISED service participants

Description: A list of the criteria which were used to determine a patient’s eligibility to be referred to the PrAISED service

### Additional File 2

Filename: Adams Additional File 2.pdf

Title: Interview schedules

Description: Interview schedules for pilot study managers, pilot study therapists, non-pilot study manager and non-pilot study therapists

