## Additional File 1 for "Implementation of the PrAISED (Promoting Activity, Independence and Stability in Early Dementia) intervention in practice: an evaluation using the Consolidated Framework for Implementation Research"

**Additional File 1. Referral criteria for PrAISED service participants**

The referral criteria for the PrAISED pilot service were:

- A diagnosis of dementia or MCI (of any subtype, except Dementia with Lewy Bodies)
- An ACE III score of 75-82/ Mini ACE of 18 to 21
- Aged 65 or over (no maximum)
- Has a family member, carer or friend who knows the participant well (defined as having contact with them for at least one hour per week via internet, telephone or in person)
- Able to walk without human help (can use walking aids)
- Capacity to give consent to participate in the service and consenting to do so
- The person with early dementia expresses an interest in participating in the service (not just the carer).

Unlike the Randomised Controlled Trial, the service was not limited to those who spoke English and there was funded access to interpreters. The service leaflets were produced in English, Urdu, Punjabi, Hindi, Polish, Ukrainian, Italian, Farsi and Spanish and translation into other languages was available.
