## Additional File 2 for "Implementation of the PrAISED (Promoting Activity, Independence and Stability in Early Dementia) intervention in practice: an evaluation using the Consolidated Framework for Implementation Research"

Interview schedules

1. Pilot Study Managers
2. Pilot Study Therapists
3. Non Pilot Study Managers
4. Non Pilot Study Therapists

### PrAISED Implementation Study

### Clinical/Research Managers (Pilot Service)

### Interview Schedule V2.0 20-10-22 Final

#### Pre-interview

My name is ……………………….., I am a ………………………. working on the PrAISED programme.

This interview is being conducted as part of the PrAISED implementation study; in this study we are aiming to carry out interviews with Clinical/Research Managers to talk about how the intervention works or might work in clinical practice.

With your agreement, we would like to record the session. The interview is strictly confidential. Your participation is voluntary and you can withdraw at any time.

Have you had a chance to read the information sheet I sent you?

Do you have any questions you would like to ask me about the research or interview?

[Complete interview Verbal or Face-to-Face Consent Form]

### Interview Questions

| A. INTRODUCTION | |
|---|---|
| **Question** | **Prompts** |
| 1. Can I ask you to introduce yourself and describe your experience of working with people with dementia? | • Can you tell me about your professional background and work at the Trust? |
| 2. What is your role in the PrAISED Pilot programme? | |
| 3. How long have you been involved in the PrAISED programme? | Were you involved in PrAISED when it was part of the research study?<br>Did your role change from what you did on the research study to the pilot programme, if yes, how, describe?<br>[If yes] *Ask questions in sections B and C<br>(if no) *Go to questions in section C |

| B. CHANGES FROM THE RESEARCH INTERVENTION TO SERVICE DELIVERY<br>***Only ask these questions to staff who were involved in the research study** | |
|---|---|
| **Question** | **Prompts** |
| ***Thinking back to PrAISED as a research study…*** | |
| 4. What changes did you need to make to delivering the PrAISED programme as a service in clinical practice? | • Referral/screening<br>• Eligibility criteria? Clinical Assessments?<br>• Content of patient letters?<br>• Therapists materials? e.g. decision tool<br>• Patient materials? e.g. Patient home file<br>• Intervention frequency, duration, session content etc. |
| 5. What were the main barriers you experienced in changing PrAISED from a research study to a service? | • Can you explain these? |
| 6. What were the main facilitators you experienced in changing PrAISED from a research study to a service? | • Can you explain these? |
| 7. How did you find the change from delivering a 12-month intervention in the research study to 3-6-months in the pilot delivery service? | • Is 3, 6, or 12 months better? Perceptions about benefits/ disadvantages for therapists/patients with longer/ shorter programme? |

| C. PILOT DELIVERY SERVICE | |
|---|---|
| **Question** | **Prompts** |
| ***I'd like to ask you about the pilot service you have been delivering….*** | |
| 8. How important do you think the PrAISED programme is for people with dementia? | • |
| 9. How well does the PrAISED programme fit with existing work processes and practices in the Trust? | • How does it work together or conflict with current programmes or processes? / In what ways do you think the PrAISED programme would replace or complement other programmes?<br>• Describe/expand on current practices of what's out there? |
| 10. How is the delivery of the PrAISED programme going so far? | • Which aspects of PrAISED do you feel are working well? What are the main facilitators?<br>• Which aspects of PrAISED do you feel are not working well? What are the main barriers? |
| 11. Has the PrAISED programme been delivered in the way you thought it would be? | • *[If Yes]* Can you describe this?<br>• *[If No]* Why not? |
| 12. Are there any specified components of the PrAISED programme that you have not been able to deliver as they were intended (as specified in the training)? | • Prompt for number, length and frequency of intervention components.<br>• *[If not]* Why not and what has been done differently? |
| 13. How did/do you or your staff use the intervention manual and materials in the pilot delivery service? | • Is/was anything else needed?<br>• Who/where did you seek help from when you had a question?<br>• Which materials were used/how were these adapted?<br>• What other materials might be needed to support intervention delivery? |
| 14. Did you put anything in place to check the quality of the programme being delivered? | • *[If Yes]* Can you describe this?<br>• *[If No]* Was there a reason for this? What could be put in place for Quality Assurance purposes? |

| | |
|---|---|
| 16. Were any changes made to the way PrAISED was delivered based on user/participant feedback? | • If yes, what changes were made and why? |
| 15. Do you think the PrAISED programme will be effective for patients referred to the service? | • What evidence do you have to support this?<br>• - e.g. feedback from patients (examples)<br>• Observed improvements in patient outcomes |

| **D. FUTURE DELIVERY OF THE PROGRAMME** | |
|---|---|
| **Question** | **Prompts** |
| ***Thinking now about the future delivery of the programme…*** | |
| 17. Are there components of the PrAISED programme that you think should be altered for use in wider clinical practice or other Trusts? | • What kinds of changes or adaptations are needed?<br>• What issues or complications might arise delivering the PrAISED programme in clinical practice? |
| 18. What else would be needed to support the commissioning of PrAISED in your Trust/other Trusts? | • Commissioning process, business case, what types of evidence? |
| 19. What would persuade you to promote commissioning of an intervention like PrAISED in the future? | • What role does research evidence/other kinds of evidence play in that decision? |
| 20. What do you think the policy drivers are in this area? | • Why do you think is it important to commission PrAISED (local/national priorities)? |
| 21. What advice would you give to another NHS Trust planning to deliver the PrAISED programme? | • |
| 22. Where would a service like PrAISED best fit in a dementia care pathway and why? | • Could you describe who/when people, along their diagnosis journey, would be referred to a service like PrAISED?<br>• What if they were discharged after an early diagnosis? Needed no other services perhaps just time to absorb their diagnosis? Who would pick this up and refer them later on? |

| | |
|---|---|
| 23. Do you think PrAISED could be delivered by other professional groups? | • *[If Yes]* Please give examples of who e.g., exercise instructors?<br>• *[If No]* Why not? |
| 24. Who trained the pilot intervention delivery staff and how were they trained?<br>25. How do you think the PrAISED training could be delivered in future? Who would deliver the training? | • Format and structure e.g. face to face or e-training/digital<br>• Content<br>• Frequency / refresher sessions<br>• Community of Practice/Network of PrAISED therapists?<br>• What else is needed in terms of training to help therapists deliver the programme? |

| **E. EXIT STRATEGY FROM PrAISED / LONG-TERM SUSTAINABILITY FOR PATIENTS** | |
|---|---|
| **Question** | **Prompts** |
| ***I would like to ask you some questions about what happens when patients complete their involvement in the PrAISED programme...*** | |
| 26. Do you have an exit strategy for the patients on completion of the PrAISED programme? | • *[If Yes]* Can you describe this? What is the purpose and scope of the strategy?<br>• *[If No]* Do you think an exit strategy could be developed? What should this involve? |
| 27. Are you aware of any dementia-friendly services that patients could be referred to after completing the PrAISED programme? | • *[If Yes]* Can you describe these? |

| **F. CLOSING QUESTIONS** | |
|---|---|
| **Question** | **Prompts** |
| 28. Is there anything we haven’t covered that you think we ought to know about PrAISED or what we have discussed today? | |

### PrAISED Implementation Study

### Clinicians/Therapists (Pilot Service)

### Interview Schedule V3.0 29-09-22

#### Pre-interview

My name is ……………………….., I am a ………………………. working on the PrAISED programme.

This interview is being conducted as part of the PrAISED implementation study; in this study we are aiming to carry out interviews with clinicians/therapists to talk about how the intervention works or might work in clinical practice.

With your agreement, we would like to record the session. The interview is strictly confidential. Your participation is voluntary and you can withdraw at any time.

Have you had a chance to read the information sheet I sent you?

Do you have any questions you would like to ask me about the research or interview?

[Complete interview Verbal or Face-to-Face Consent Form]

### Interview Questions

| A. INTRODUCTION | |
|---|---|
| **Question** | **Prompts** |
| 29. Can I ask you to introduce yourself and describe your experience of working with people with dementia? | • Can you tell me about your professional background and work at the Trust? |
| 30. What is your role in the PrAISED programme? | • How long have you been involved in the PrAISED programme? |
| 31. Were you involved in PrAISED when it was part of the research study?<br><br>[If yes *Ask questions in section B]<br><br>[If no, go to section C] | • |

| B. CHANGE FROM RESEARCH INTERVENTION TO PILOT SERVICE<br>*Only ask these questions to staff who were involved in the research study | |
|---|---|
| **Question** | **Prompts** |
| ***I'd like to ask you about the research intervention and pilot service you have been delivering….*** | |
| 32. What changes were made to the PrAISED programme in order to deliver it as a service in clinical practice instead of a research study? | [May need to distinguish here between changes that were made by the team to the overall programme vs individual changes that were made in the way they delivered their sessions]<br>• Referral/screening processes<br>• Eligibility criteria? Clinical Assessments?<br>• Content of patient letters?<br>• Therapists materials? e.g. decision tool<br>• Patient materials? e.g. Patient home file<br>• Intervention frequency, duration, session content etc.<br>• Existing paperwork/computer systems? |
| 33. What were the main barriers you experienced in changing PrAISED from a research study to a service? | • Can you explain these? |
| 34. What were the main facilitators you experienced in changing PrAISED from a research study to a service? | • Can you explain these? |
| 35. How did you find changing from delivering a 12-month intervention in the research study to a 3-6-month study in the pilot delivery service? | • Is 3, 6 or 12 months better? Perceptions about benefits/ disadvantages for therapists/patients with longer/ shorter programme? |

| C. PILOT DELIVERY SERVICE | |
|---|---|
| **Question** | **Prompts** |
| ***I'd like to ask you about the pilot service you have been delivering….*** | |
| 36. How do you feel about the PrAISED programme being delivered in your setting as a routine service? | • How well does the PrAISED programme fit with existing work processes and practices in the Trust?<br>• How does it complement or conflict with current programmes or processes?<br>• Could it replace other programmes? |
| 37. How is the delivery of the PrAISED service going so far? | • Which aspects of PrAISED do you feel are working well? What are the main facilitators? Why?<br>• Which aspects of PrAISED do you feel are not working well? What are the main barriers? Why? |
| 38. Has the PrAISED pilot service been delivered in the way you thought it would be? | • *[If Yes]* Can you describe this?<br>• *[If No]* Why not? |
| 39. Are there any specified components of the PrAISED Service that you have not been able to deliver as they were intended (as specified in the training), or that you have delivered differently? | • Prompt for number, length, frequency and content of intervention components.<br>• *[If not]* Why not? |
| 40. How did/do you use the intervention manual and materials in the pilot delivery service? | • Is/was anything else needed?<br>• Who/where did you seek help from when you had a question?<br>• Which materials were used/how were these adapted?<br>• What other materials might be needed to support intervention delivery? |
| 41. Do you think the PrAISED programme will be effective for patients referred to the service? | • What evidence do you have to support this?<br>• - e.g. feedback from patients (examples)<br>• Observed improvements in patient outcomes |

| D. PARTICIPANT RESPONSE /INVOLVEMENT | |
|---|---|
| **Question** | **Prompts** |
| 42. How would you describe participant response to the PrAISED programme? | • For example, indicators such as levels of participation and enthusiasm / completion of the programme<br>• What enablers have supported patients participating in the PrAISED programme?<br>• What barriers have patients faced participating in the PrAISED programme? |
| 43. Were any changes made to the way PrAISED was delivered based on user feedback? | • If yes, what changes were made and why? |

| E. FUTURE DELIVERY OF THE PROGRAMME | |
|---|---|
| **Question** | **Prompts** |
| ***Thinking now about the future delivery of the programme…*** | |
| 44. Are there components of the PrAISED programme that you think should be altered for use in wider clinical practice or other Trusts? | • What kinds of changes or adaptations are needed?<br>• What issues or complications might arise delivering the PrAISED programme in clinical practice? |
| 45. What advice would you give to another therapist planning to deliver the PrAISED programme? | |
| 46. What skills/experience/qualifications do you think someone needs to undertake the role of therapist/RSW on PrAISED? | • Do you think PrAISED could be delivered by other professional groups?<br>*[If Yes]* Please give examples of who e.g., exercise instructors?<br>*[If No]* Why not? |

| F. THERAPISTS' SELF EFFICACY | |
|---|---|
| **Question** | **Prompts** |
| 47. How have you found the training you received in PrAISED (e.g., initial training and ongoing support)? | • Did you feel you had enough training to effectively deliver the programme?<br>• How confident do you feel in your professional role to deliver the PrAISED programme?<br>• |
| 48. What or who do you think would be suitable to deliver the PrAISED training in future? | • Format and structure e.g. face to face or e-training/digital<br>• Content<br>• Frequency / refresher sessions<br>• Community of Practice/Network of PrAISED therapists?<br>• What else is needed in terms of training and support to help therapists deliver the programme? |

| G. EXIT STRATEGY FROM PrAISED / LONG-TERM SUSTAINABILITY FOR PATIENTS | |
|---|---|
| **Question** | **Prompts** |
| ***Finally, I would like to ask you some questions about what happens when patients complete their involvement in the PrAISED programme...*** | |
| 49. Do you have an exit strategy / discharge plan for the patients on completion of the PrAISED programme? | • *[If Yes]* Can you describe this? What is the purpose and scope of the strategy?<br>• *[If No]* Do you think an exit strategy could be developed? What should this involve? |
| 50. What advice or information do you offer patients during or at the end of the PrAISED programme to support them in maintaining their activity and health? | • What, when, where?<br>• Signposting or referral?<br>• Resources available? |
| 51. Are you aware of any dementia-friendly services that patients could be referred to after completing the PrAISED programme? | • *[If Yes]* Can you describe these?<br>• How might these compete with or complement PrAISED? |

| H. CLOSING QUESTIONS | |
|---|---|
| **Question** | **Prompts** |
| 52. Is there anything we haven’t covered that you think we ought to know about PrAISED or what we have discussed today? | |

### PrAISED Implementation Study

### Clinical/Research Managers (Non-Pilot Service Sites)

### Interview Schedule V2.0 30-06-22

#### Pre-interview

My name is ……………………….., I am a ………………………. working on the PrAISED programme.

This interview is being conducted as part of the PrAISED implementation study; in this study we are aiming to carry out interviews with Clinical/Research Managers to talk about how the intervention works or might work in clinical practice.

With your agreement, we would like to record the session. The interview is strictly confidential. Your participation is voluntary and you can withdraw at any time.

Have you had a chance to read the information sheet I sent you?

Do you have any questions you would like to ask me about the research or interview?

[Complete interview Verbal or Face-to-Face Consent Form]

### Interview Questions

| A. INTRODUCTION | |
|---|---|
| **Question** | **Prompts** |
| 53. Can I ask you to introduce yourself and describe your experience of working with people with dementia? | • Can you tell me about your professional background and work at the Trust? |
| 54. What is your role in the PrAISED programme? | • How long were you involved in the PrAISED research programme for? |

| B. FUTURE DELIVERY OF THE PROGRAMME | |
|---|---|
| **Question** | **Prompts** |
| ***Thinking about the future delivery of the programme…*** | |
| ***55.*** How would you feel about the PrAISED programme being delivered in your setting as a routine service? | • How well does the PrAISED programme fit with existing work processes and practices in the Trust?<br> ◦ How does it work together or conflict with current programmes or processes? |
| 56. In what ways do you think the PrAISED programme would replace or complement other programmes? | |
| 57. What changes do you think would need to be made in order to deliver the PrAISED programme as a service in clinical practice instead of a research study? | • Referral/screening processes<br>• Eligibility criteria? Clinical Assessments?<br>• Content of patient letters?<br>• Therapists materials? e.g. decision tool<br>• Patient materials? e.g. Patient home file<br>• Intervention frequency, duration, session content etc. |
| 58. Are there components of the PrAISED programme that you think should be altered for use in wider clinical practice or other Trusts? | • What kinds of changes or adaptations are needed? |
| 59. What do you think would be the main barriers in delivering the PrAISED programme in routine clinical practice? | • Can you explain these? |
| 60. What would help to facilitate delivering the PrAISED programme in routine clinical practice? | • Can you explain these? |
| 61. What might you put in place to check the quality of the programme being delivered? | • Quality assurance processes |
| 62. What advice would you give to another NHS Trust planning to deliver the PrAISED programme? | • |
| 63. Do you think PrAISED could be delivered by other professional groups? | • *[If Yes]* Please give examples e.g., exercise instructors?<br>• *[If No]* Why not? |
| 64. How do you think the PrAISED training could be delivered in future? | • Format and structure e.g. face to face or e-training/digital<br>• Content |

| | |
|---|---|
| | • Frequency / refresher sessions<br>• Community of Practice/Network of PrAISED therapists?<br>• What else is needed in terms of training or support to help therapists deliver the programme? |
| ***Thinking about the future of the funding / commissioning of the PrAISED programme…*** | |
| 65. What would be needed to support the commissioning of PrAISED in your Trust/other Trusts? | • Commissioning process, business case, what types of evidence? |
| 66. What would persuade you to promote commissioning of an intervention like PrAISED in the future? | • What role does research evidence/other kinds of evidence play in that decision? |
| 67. What do you think the policy drivers are in this area / field? | • Why do you think is it important to commission PrAISED (local/national priorities)? |
| 68. Who do you think *could* fund the PrAISED the intervention? | |
| 69. Who do you think *would* fund the intervention? | |
| 70. How could delivery of the intervention delivery/funding be sustained? | |

| **C. EXIT STRATEGY FROM PrAISED / LONG-TERM SUSTAINABILITY FOR PATIENTS** | |
|---|---|
| **Question** | **Prompts** |
| ***I would like to ask you about what happens when patients complete their involvement in the PrAISED programme...*** | |
| 71. Did you have an exit strategy for the patients on completion of the PrAISED programme? | • *[If Yes]* Can you describe this? What is the purpose and scope of the strategy?<br>• What advice or information was offered to patients?<br>• *[If No]* Do you think an exit strategy could be developed? What should this involve? |
| 72. Are you aware of any dementia-friendly services that patients could be referred to after completing the PrAISED programme? | • *[If Yes]* Can you describe these? |

| D. CLOSING QUESTIONS | |
|---|---|
| **Question** | **Prompts** |
| 73. Is there anything we haven't covered that you think we ought to know about PrAISED or what we have discussed today? | |

### PrAISED Implementation Study

### Clinicians/Therapists (Non-Pilot Service Sites)

### Interview Schedule V2.0 18-07-22

#### Pre-interview

My name is ……………………….., I am a ………………………. working on the PrAISED programme.

This interview is being conducted as part of the PrAISED implementation study; in this study we are aiming to carry out interviews with clinicians/therapists to talk about how the intervention works or might work in clinical practice.

With your agreement, we would like to record the session. The interview is strictly confidential. Your participation is voluntary and you can withdraw at any time.

Have you had a chance to read the information sheet I sent you?

Do you have any questions you would like to ask me about the research or interview?

[Complete interview Verbal or Face-to-Face Consent Form]

### Interview Questions

| A. INTRODUCTION | |
|---|---|
| **Question** | **Prompts** |
| 74. Can I ask you to introduce yourself and describe your experience of working with people with dementia? | • Can you tell me about your professional background and work at the Trust? |
| 75. What was your role in the PrAISED programme? | • How long were you involved in the PrAISED research programme for? |

| B. FUTURE DELIVERY OF THE PROGRAMME | |
|---|---|
| **Question** | **Prompts** |
| ***Thinking about the future delivery of the programme…*** | |
| ***76.*** How would you feel about the PrAISED programme being delivered in your setting as a routine service? | • How well does the PrAISED programme fit with existing work processes and practices in the Trust?<br>• How would it work together or conflict with current programmes or processes? |
| 77. What changes do you think would need to be made in order to deliver the PrAISED programme as a service in clinical practice instead of a research study? | • Referral/screening processes<br>• Eligibility criteria? Clinical Assessments?<br>• Content of patient letters?<br>• Therapists materials? e.g. decision tool<br>• Patient materials? e.g. Patient home file<br>• Intervention frequency, duration, session content etc. |
| 78. What do you think would be the main barriers in delivering the PrAISED programme in routine clinical practice? | • Can you explain these? |
| 79. What would help to facilitate delivering the PrAISED programme in routine clinical practice? | • Can you explain these? |
| 80. What advice would you give to another therapist planning to deliver the PrAISED programme? | |
| 81. In what ways do you think the PrAISED programme would replace or complement other programmes? | |
| 82. What skills/experience/qualifications do you think someone needs to undertake the role of therapist/RSW on PrAISED? | • Do you think PrAISED could be delivered by other professional groups?<br>*[If Yes]* Please give examples of who e.g., exercise instructors?<br>*[If No]* Why not? |

| C. THERAPISTS' SELF EFFICACY | |
|---|---|
| **Question** | **Prompts** |
| 83. How did you find the training you received in PrAISED (e.g., initial training and ongoing support)? | • Did you feel you had enough training to effectively deliver the programme?<br>• How confident did you feel in your professional role to deliver the PrAISED programme? |
| 84. What else is needed in terms of training and support to help therapists deliver the programme? | |
| 85. What do you think is needed to deliver the PrAISED training in future? | • Format and structure e.g., face to face or e-training/digital<br>• Content<br>• Frequency / refresher sessions<br>• Community of Practice/Network of PrAISED therapists? |

| D. EXIT STRATEGY FROM PrAISED / LONG-TERM SUSTAINABILITY FOR PATIENTS | |
|---|---|
| **Question** | **Prompts** |
| ***I would like to ask you some questions about what happens when patients complete their involvement in the PrAISED programme...*** | |
| 86. Did you have an exit strategy / discharge plan for the patients on completion of the PrAISED programme? | • *[If Yes]* Can you describe this? What is the purpose and scope of the strategy?<br>• *[If No]* Do you think an exit strategy could be developed? What should this involve? |
| 87. What advice or information did you offer patients during or at the end of the PrAISED programme to support them in maintaining their activity and health? | • What, when, where?<br>• Signposting or referral? |
| 88. Are you aware of any dementia-friendly services that patients could be referred to after completing the PrAISED programme? | • *[If Yes]* Can you describe these?<br>• How might these compete with or complement PrAISED? |

| E. CLOSING QUESTIONS | |
|---|---|
| **Question** | **Prompts** |
| 89. Is there anything we haven’t covered that you think we ought to know about PrAISED or what we have discussed today? | |
